# Evaluation of cerebrospinal fluid alpha-synuclein seed amplification assay in PSP and CBS

**DOI:** 10.1101/2024.02.28.24303478

**Authors:** DP Vaughan, R Fumi, M Theilmann Jensen, T Georgiades, L Wu, D Lux, R Obrocki, J Lamoureux, O Ansorge, KSJ Allinson, TT Warner, Z Jaunmuktane, A Misbahuddin, PN Leigh, BCP Ghosh, KP Bhatia, A Church, C Kobylecki, MTM Hu, JB Rowe, C Blauwendraat, HR Morris, E Jabbari

## Abstract

**Background:** Seed amplification assay (SAA) testing has become an important biomarker in the diagnosis of alpha-synuclein related neurodegenerative disorders.

**Objectives:** To assess the rate of alpha-synuclein SAA positivity in progressive supranuclear palsy (PSP) and corticobasal syndrome (CBS), and analyse the clinical and pathological features of SAA positive and negative cases.

**Methods:** 106 CSF samples from clinically diagnosed PSP (n=59), CBS (n=37) and indeterminate parkinsonism cases (n=10) were analysed using alpha-synuclein SAA.

**Results:** Three cases (1 PSP, 2 CBS) were Multiple System Atrophy (MSA)-type SAA positive. 5/59 (8.5%) PSP cases were Parkinson’s disease (PD)-type SAA positive, and these cases were older and had a shorter disease duration compared with SAA negative cases. In contrast, 9/35 (25.7%) CBS cases were PD-type SAA positive.

**Conclusions:** Our results suggest that PD-type seeds can be detected in PSP and CBS using a CSF alpha-synuclein SAA, and in PSP this may impact on clinical course.

## Introduction

Progressive supranuclear palsy (PSP) and Corticobasal syndrome (CBS) are Parkinson-plus disorders with associated eye movement and cognitive impairments, and have an estimated prevalence of 3-10 per 100,000 **(1)**. The neuropathology of PSP involves accumulation of neuronal and glial hyperphosphorylated 4-repeat tau pathology with associated neurodegeneration **(2)**, and a clinical diagnosis of PSP is strongly predictive of underlying PSP pathology at post-mortem **(3)**. In contrast, the underlying neuropathology of CBS is diverse including corticobasal degeneration (also a 4-repeat tauopathy, CBD), PSP and Alzheimer’s disease (AD) **(4)**.

Despite recent updates to clinical diagnostic criteria for PSP and CBS which acknowledge the clinical heterogeneity of these conditions **(5) (6)**, the diagnosis of PSP and CBS in the early stages of symptomatic disease is challenging. This is due to clinical overlap with other parkinsonian disorders including Parkinson’s disease (PD), Dementia with Lewy bodies (DLB) and multiple system atrophy (MSA) which are characterised by the accumulation of alpha-synuclein neuropathology, and the lack of objective disease-specific diagnostic biomarkers for all parkinsonian disorders. This affects the recruitment of early-stage patients for clinical trials, while misdiagnosis reduces the power of tau-targeting trials.

There is increasing evidence that alpha-synuclein seed amplification assays (SAAs) can reliably differentiate people with PD from healthy controls with high sensitivity and specificity, and may even be able to differentiate PD from other synucleinopathies including MSA **(7)**. Additionally, the assay results show good inter-laboratory concordance **(8)**, can be applied to a range of biofluids (including CSF, blood and saliva) with variable sensitivities and specificities, and show a high rate of positivity in the prodromal phase of synucleinopathies **(7)**. Consequently, there is a drive to move away from a clinical diagnosis of PD towards a biological definition of neuronal alpha-synuclein disease based on SAA positivity to enable clinical trials at early disease stages **(9)**. SAAs are based on the self-templating abilities of pathologically misfolded proteins, including alpha-synuclein and tau, to aggregate and induce amyloid fibril formation and so there are ongoing efforts to develop effective 4-repeat tau SAAs **(10)**.

Here, we evaluate the alpha-synuclein SAA in a cohort of PSP and CBS CSF samples with matched clinical and post-mortem data, and discuss the implications of these results.

## Methods

### Participant recruitment and diagnosis

Written informed consent was obtained from people with PSP and CBS who were recruited to the PROSPECT-UK study between September 1, 2015 to November 1, 2023 **(Supplementary information)**. Participants were either retrospectively or prospectively assigned a clinical diagnosis (+/- associated subtype) in line with the 2017 MDS-PSP diagnostic criteria and 2013 Armstrong CBS diagnostic criteria **(5) (6)**, achieving “possible” or “probable” criteria. Of note, participants who fulfilled criteria for a PSP/CBS overlap syndrome were included in the PSP group for analysis. We also included indeterminate (IDT) parkinsonism participants who were suspected to have a Parkinson-plus disorder but did not fulfil PSP or CBS diagnostic criteria.

### Clinical and neuropathological data collection

We completed core clinical assessments at baseline, and where possible these were repeated after 6, 12, 24, and 36 months of follow-up. At the baseline visit we obtained clinical data including sex, age at symptom onset, age and disease duration at baseline assessment, and clinician diagnostic certainty (ranked 0-100). At each study visit, we obtained scores for the PSP Rating Scale (PSPRS), the MDS Unified Parkinson’s Disease Rating Scale (MDS-UPDRS), the Montreal Cognitive Assessment (MoCA), and the Schwab and England Activities of Daily Living Scale (SEADL). Olfactory function was assessed in an unbiased subset of participants using the “Sniffin’ Sticks” test with hyposmia defined as a score < 11 out of 16.

Lumbar puncture was an optional study assessment to obtain baseline CSF for biomarker analyses. Mortality data was censored on January 1, 2024 and in deceased cases a total disease duration from symptom onset to death was calculated. In those undergoing post-mortem, the primary neuropathological diagnosis was noted along with the presence or absence of Lewy body co-pathology and associated Braak stage.

Baseline CSF samples from participants were applied to the Amprion alpha-synuclein SAA **(Supplementary information)**. Kinetic parameters of the aggregation curve were used to assign positive results as “PD-type” or “MSA-type” as described previously **(11)**.

### Statistical analysis

Group comparisons were performed with a χ2 test for categorical clinical variables. T-test and logistic regression, adjusting for sex and age at assessment, were used for group comparisons of continuous clinical variables. Statistical significance was set at *p <* 0.05. All statistical analyses were performed in GraphPad Prism 9.1.1 (San Diego, CA).

### Data availability

All data used in this study will be released via GP2 and can be accessed at https://www.amp-pd.org/.

## Results

106 participants in the PROSPECT-UK study who had undergone a baseline lumbar puncture and clinical assessment were included in this study consisting of 59 clinically diagnosed PSP participants, 37 clinically diagnosed CBS participants and 10 IDT participants. Aside from differing sex distributions, the baseline clinical characteristics of the PSP and CBS groups were largely comparable and are summarised in **Table 1**. In those undergoing post-mortem analysis (n=9), an ante-mortem clinical diagnosis of PSP was always predictive of underlying 4-repeat tau neuropathology (4 PSP; 1 CBD), in contrast to CBS (1 PSP; 1 CBD; 1 MSA; 1 AD).

**Table 1:** Clinical characteristics of PSP and CBS groups.

|  | **PSP (n=59)** | **CBS (n=37)** | **IDT (n=10)** |
| --- | --- | --- | --- |
| **Sex (% male)** | 58% | 30% * | 30% |
| **Alpha-synuclein SAA status** | Negative, n=52<br>PD-type, n=5<br>MSA-type, n=1<br>Indeterminate, n=1 | Negative, n=26<br>PD-type, n=9<br>MSA-type, n=2 | Negative, n=9<br>PD-type, n=1 |
| **Clinical subtype at baseline assessment (n)** | PSP-RS = 33<br>PSP-P = 11<br>PSP-PGF = 6<br>PSP/CBS = 5<br>PSP-SL = 2<br>PSP-F = 2 |  |  |
| **Primary pathological diagnosis in participants undergoing post-mortem (n)** | PSP, n=4<br>CBD, n=1 | CBD, n=1<br>PSP, n=1<br>MSA, n=1<br>AD, n=1 | N/A |
| **Diagnostic certainty at baseline assessment (0-100), median (range)** | 85 (50-100) | 75 (50-95) | 60 (45-60) |
| **Mean age at symptom onset, years (SD)** | 65.0 years (6.9) | 63.5 years (7.5) | 68.3 years (5.6) |
| **Mean age at baseline assessment, years (SD)** | 69.0 years (6.8) | 68.2 years (7.5) | 72.1 years (5.6) |
| **Mean disease duration at baseline assessment, years (SD)** | 4.0 years (2.0) | 4.7 years (2.1) | 3.8 years (1.2) |
| **Mean PSPRS at baseline assessment, score (SD)** | 33.7 (15.0) | 31.0 (12.0) | 36.5 (12.8) |
| **Mean MDS-UPDRS III at baseline assessment, score (SD)** | 36.3 (14.5) | 43.9 (14.0) | 37.5 (15.8) |
| **Mean MoCA at baseline assessment, score (SD)** | 22.2 (4.8) | 19.6 (5.8) | 19.6 (6.3) |
| **Median SEADL at baseline assessment, score (range)** | 60 (10-90) | 50 (20-90) | 55 (20-60) |
| **Participants undergoing assessment at 2 or more time-points (n), mean latency between baseline and final assessments, years (SD)** | n=34<br>1.6 years (0.9) | n=17<br>1.7 years (0.7) | n=2<br>1.2 years (0.2) |
| **Participants deceased at censoring (n), mean disease duration from symptom onset to death, years (SD)** | n=28<br>7.0 years (3.0) | n=19<br>8.2 years (3.2) | n=5<br>6.2 years (2.9) |

Alzheimer's disease: AD, Corticobasal syndrome: CBS, Corticobasal degeneration: CBD, Indeterminate: IDT, Multiple system atrophy: MSA, Progressive Supranuclear Palsy: PSP, PSP-Richardson syndrome: PSP-RS, PSP-parkinsonism: PSP-P, PSP-progressive gait freezing: PSP-PGF, PSP/CBS overlap: PSP/CBS, PSP-speech and language disorder: PSP-SL, PSP-frontal: PSP-F, number: n, standard deviation: SD, PSP rating scale: PSPRS, Movement Disorder Society-Unified Parkinson's Disease Rating Scale part III: MDS-UPDRS III, Montreal Cognitive Assessment: MoCA, Schwab and England activities of daily living scale: SEADL. \* = P<0.05 vs. PSP group using $\chi^2$ test.

5/59 (8.5%) PSP participants were PD-type alpha-synuclein SAA positive, one participant had an indeterminate result, and one participant was MSA-type positive. In contrast, 9/37 (24.3%) CBS participants were PD-type alpha-synuclein SAA positive and two participants were MSA-type positive, one of which had post-mortem confirmation of MSA as the primary pathological diagnosis. Of the total cohort of post-mortem cases, only 1 case (AD pathological diagnosis) was PD-type SAA positive, and this case had Braak stage 1 Lewy body co-pathology. In the remaining post-mortem cases, one PSP case had Braak stage 1 Lewy body co-pathology but was PD-type SAA negative.

We then divided the PSP and CBS groups into PD-type SAA positive and negative groups. Alongside looking for differences in clinical and pathological variables described above, we also collected data on: 1) change in clinical diagnosis since the baseline assessment; 2) presence or absence of tremor, postural lightheadedness and visual hallucinations from the MDS-UPDRS parts I and II, and hyposmia as per the “Sniffin’ Sticks” test, as these features are frequently encountered in synucleinopathies (PD, MSA and DLB) but are rarely seen in 4-repeat tauopathies.

Although underpowered to detect statistical significance, this exploratory analysis showed a trend towards SAA positive PSP participants being older, more impaired in motor, cognitive and functional scales, and having a more aggressive rate of progression compared with SAA negative PSP participants. In those undergoing serial clinical assessment, change in diagnosis in the PSP group was rare (2/33, 6.1%) and the rates of synucleinopathy clinical features were similar in SAA positive and negative groups with the exception of hyposmia which was more marked in the SAA positive group. In contrast, SAA positive vs. negative clinical differences in the CBS group were less marked, although this was in the context of a higher overall rate of change in diagnosis (2/16, 12.5%) and long disease duration in SAA positive cases who were still alive at the point of censoring **(Table 2)**.

**Table 2:**
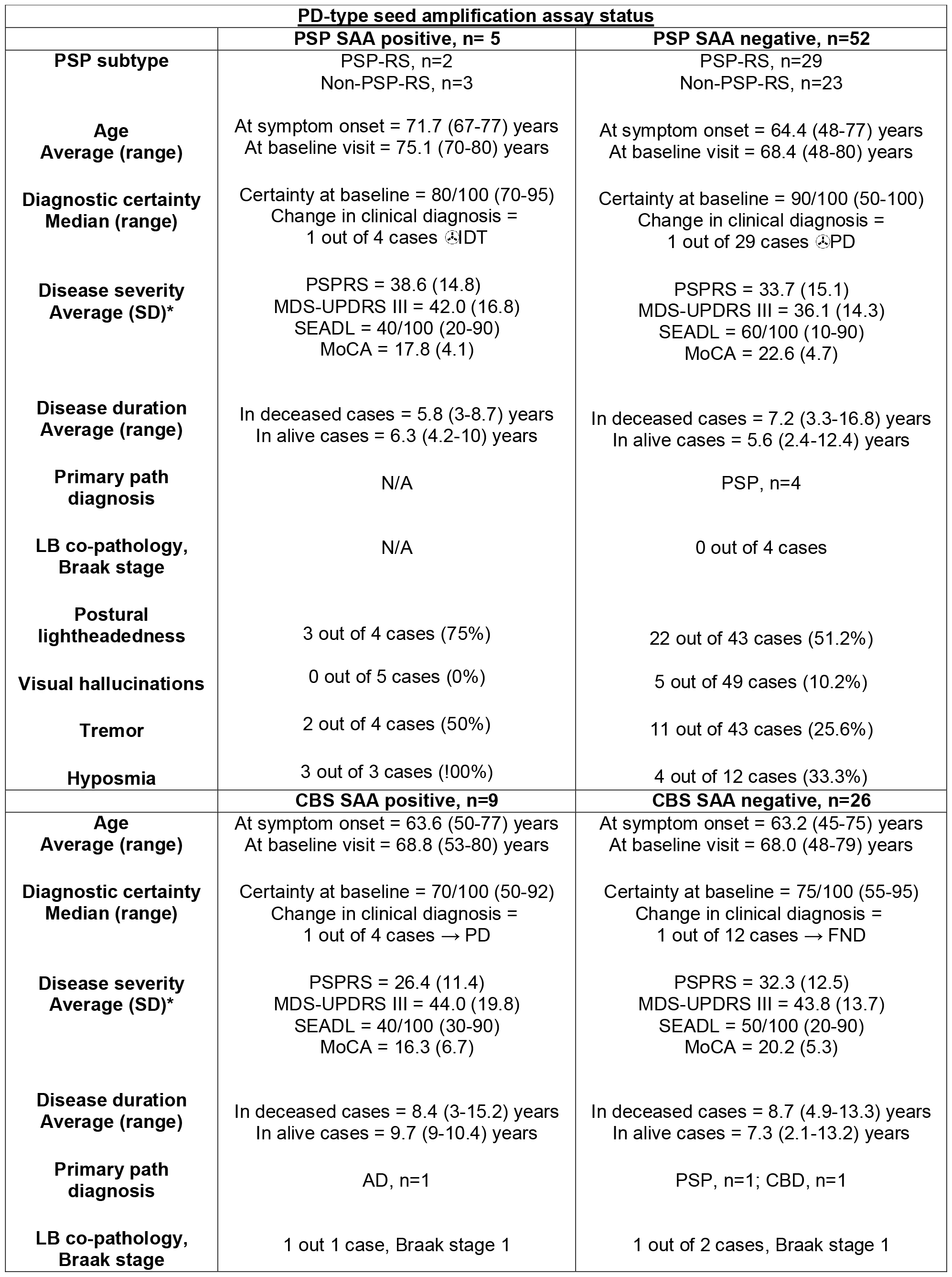
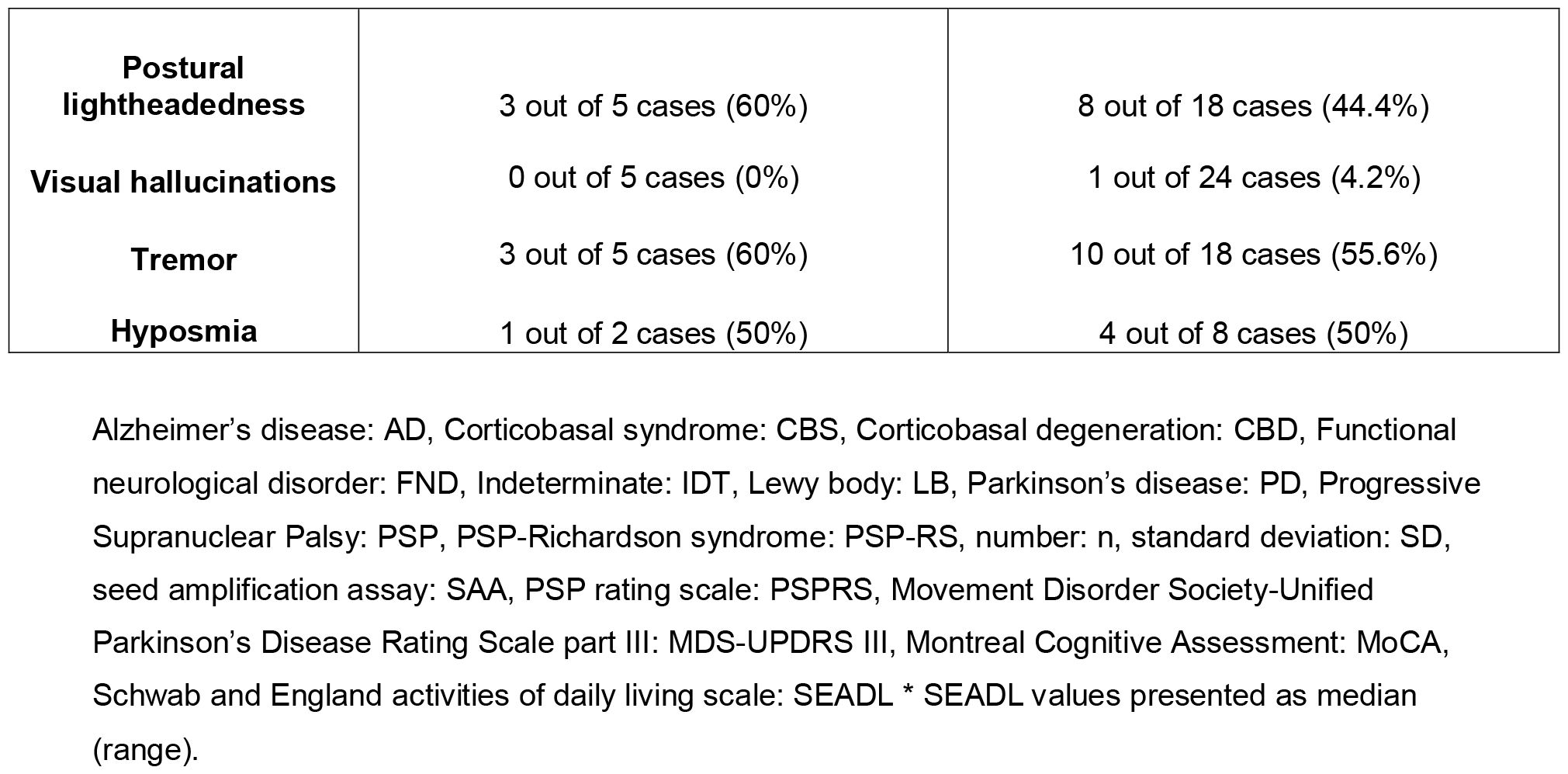
PD-type SAA status in PSP and CBS groups.

| **PD-type seed amplification assay status** |  |  |
| --- | --- | --- |
|  | **PSP SAA positive, n= 5** | **PSP SAA negative, n=52** |
| **PSP subtype** | PSP-RS, n=2<br>Non-PSP-RS, n=3 | PSP-RS, n=29<br>Non-PSP-RS, n=23 |
| **Age**<br>**Average (range)** | At symptom onset = 71.7 (67-77) years<br>At baseline visit = 75.1 (70-80) years | At symptom onset = 64.4 (48-77) years<br>At baseline visit = 68.4 (48-80) years |
| **Diagnostic certainty**<br>**Median (range)** | Certainty at baseline = 80/100 (70-95)<br>Change in clinical diagnosis =<br>1 out of 4 cases → IDT | Certainty at baseline = 90/100 (50-100)<br>Change in clinical diagnosis =<br>1 out of 29 cases → PD |
| **Disease severity**<br>**Average (SD)\*** | PSPRS = 38.6 (14.8)<br>MDS-UPDRS III = 42.0 (16.8)<br>SEADL = 40/100 (20-90)<br>MoCA = 17.8 (4.1) | PSPRS = 33.7 (15.1)<br>MDS-UPDRS III = 36.1 (14.3)<br>SEADL = 60/100 (10-90)<br>MoCA = 22.6 (4.7) |
| **Disease duration**<br>**Average (range)** | In deceased cases = 5.8 (3-8.7) years<br>In alive cases = 6.3 (4.2-10) years | In deceased cases = 7.2 (3.3-16.8) years<br>In alive cases = 5.6 (2.4-12.4) years |
| **Primary path**<br>**diagnosis** | N/A | PSP, n=4 |
| **LB co-pathology,**<br>**Braak stage** | N/A | 0 out of 4 cases |
| **Postural**<br>**lightheadedness** | 3 out of 4 cases (75%) | 22 out of 43 cases (51.2%) |
| **Visual hallucinations** | 0 out of 5 cases (0%) | 5 out of 49 cases (10.2%) |
| **Tremor** | 2 out of 4 cases (50%) | 11 out of 43 cases (25.6%) |
| **Hyposmia** | 3 out of 3 cases (100%) | 4 out of 12 cases (33.3%) |
|  | **CBS SAA positive, n=9** | **CBS SAA negative, n=26** |
| **Age**<br>**Average (range)** | At symptom onset = 63.6 (50-77) years<br>At baseline visit = 68.8 (53-80) years | At symptom onset = 63.2 (45-75) years<br>At baseline visit = 68.0 (48-79) years |
| **Diagnostic certainty**<br>**Median (range)** | Certainty at baseline = 70/100 (50-92)<br>Change in clinical diagnosis =<br>1 out of 4 cases → PD | Certainty at baseline = 75/100 (55-95)<br>Change in clinical diagnosis =<br>1 out of 12 cases → FND |
| **Disease severity**<br>**Average (SD)\*** | PSPRS = 26.4 (11.4)<br>MDS-UPDRS III = 44.0 (19.8)<br>SEADL = 40/100 (30-90)<br>MoCA = 16.3 (6.7) | PSPRS = 32.3 (12.5)<br>MDS-UPDRS III = 43.8 (13.7)<br>SEADL = 50/100 (20-90)<br>MoCA = 20.2 (5.3) |
| **Disease duration**<br>**Average (range)** | In deceased cases = 8.4 (3-15.2) years<br>In alive cases = 9.7 (9-10.4) years | In deceased cases = 8.7 (4.9-13.3) years<br>In alive cases = 7.3 (2.1-13.2) years |
| **Primary path**<br>**diagnosis** | AD, n=1 | PSP, n=1; CBD, n=1 |
| **LB co-pathology,**<br>**Braak stage** | 1 out 1 case, Braak stage 1 | 1 out of 2 cases, Braak stage 1 |
| **Postural lightheadedness** | 3 out of 5 cases (60%) | 8 out of 18 cases (44.4%) |
| **Visual hallucinations** | 0 out of 5 cases (0%) | 1 out of 24 cases (4.2%) |
| **Tremor** | 3 out of 5 cases (60%) | 10 out of 18 cases (55.6%) |
| **Hyposmia** | 1 out of 2 cases (50%) | 4 out of 8 cases (50%) |

Alzheimer's disease: AD, Corticobasal syndrome: CBS, Corticobasal degeneration: CBD, Functional neurological disorder: FND, Indeterminate: IDT, Lewy body: LB, Parkinson's disease: PD, Progressive Supranuclear Palsy: PSP, PSP-Richardson syndrome: PSP-RS, number: n, standard deviation: SD, seed amplification assay: SAA, PSP rating scale: PSPRS, Movement Disorder Society-Unified Parkinson's Disease Rating Scale part III: MDS-UPDRS III, Montreal Cognitive Assessment: MoCA, Schwab and England activities of daily living scale: SEADL \* SEADL values presented as median (range).

## Discussion

In this study we have detected alpha-synuclein SAA positivity in people with a clinical diagnosis of PSP and CBS, and have considered the clinico-pathological implications of this.

The three MSA-type SAA positive cases (one of which had post-mortem confirmation) highlight the benefit of having disease-specific biomarkers for Parkinson-plus disorders. In our PSP group, we found low rates of change in clinical diagnosis with follow-up and high rates of primary 4-repeat tau pathology in cases that came to post-mortem which is consistent with previous studies **(3)**, and so PD-type SAA positivity is highly unlikely to represent cases with primary PD pathology. Indeed, our PSP group rate of 5/59 (8.5%) PD-type SAA positivity is higher than the background positivity rate (∼4%) seen in similar-aged neurologically normal controls **(7)**, and is comparable with the rates of Lewy body co-pathology seen in previous post-mortem studies of PSP **(12) (13) (14)** as well as the MD-GAP post-mortem study of UK brain banks which found Lewy body co-pathology in 43/740 (5.8%) of PSP cases (unpublished data). In contrast, a recent study found PD-type SAA positivity in 4/16 (25%) PSP CSF samples **(15)**. Of interest, our data suggest that PD co-pathology may be seen in older PSP patients, which may suggest that age-related impairment in proteostasis leads to the accumulation of multiple co-pathologies **(12)**. Additionally, in contrast with previous clinicopathological studies **(13) (14)**, our data provides preliminary evidence that PD co-pathology may increase disease severity and the rate of disease progression which would have implications for clinical trial design.

PD-type SAA positivity was relatively high in our CBS group (9/37, 24.3%). Cortical Lewy body disease can present with corticobasal syndrome, but previous studies suggest that this is a rare finding, so it is unlikely that a quarter of our CBS cases had primary Lewy body pathology **(16) (17)**. Our SAA positivity rates were higher than the rates of Lewy body co-pathology observed in CBD in previous neuropathology studies (10%) **(12)** and in the MD-GAP database 13/142 (9.2%) (unpublished data). This raises the possibility that our higher rates of SAA positivity may be due to the CBS group containing a mixture of cases who have been clinically misdiagnosed, i.e. have primary PD pathology, and cases who have underlying primary 4-repeat tau pathology (PSP or CBD) and Lewy body co-pathology. Alternatively, it may represent poor specificity of the alpha-synuclein SAA in CBD cases. Conversely, our Lewy body co-pathology rates may be underestimated as the sensitivity of alpha-synuclein SAAs are limited in low Braak stages of Lewy body pathology **(18)**.

Despite important overall conclusions, our study has limitations mainly around power and the potential for inter-rater variability to impact on our clinical measures. As well as replication of our findings in better-powered cohorts with higher rates of matched post-mortem data, important follow-up work will be to combine the testing of alpha-synuclein and 4-repeat tau SAAs across multiple parkinsonian disorders to assess positive and negative predictive values of both assays. It will also be important to assess fluid biomarker and neuropathological correlates of AD co-pathology, its impact on disease progression, and the association with common genetic variants of interest including *MAPT* and *APOE4* status.

## Acknowledgements

We thank the patients and their families for study participation. The PROSPECT-UK study is supported by the PSP Association, CBD Solutions and the Medical Research Council UK. This work was supported in part by the Intramural Research Program of the National Institutes of Health including: the Center for Alzheimer’s and Related Dementias, within the Intramural Research Program of the National Institute on Aging and the National Institute of Neurological Disorders and Stroke.

## Financial disclosures and competing interests

As below.

### Contributions

HRM is the chief investigator of the PROSPECT-UK study; DV, RF, MTJ, DL, RO, AM, PNL, KPB, BCPG, AC, CK, MTMH, JBR, HRM and EJ carried out PROSPECT-UK study assessments; OA, KSJA, TTW and ZJ carried out post-mortem assessments; DV, RF, MTJ, TG, LW, JL, CB and EJ contributed to the acquisition and analysis of data; EJ conceptualised and supervised the study; DV and EJ drafted the manuscript; all authors critically reviewed the manuscript.

