## Supplementary information for "Evaluation of cerebrospinal fluid alpha-synuclein seed amplification assay in PSP and CBS"

**Participant recruitment**

People with PSP and CBS who were recruited to the UK-wide PROSPECT study natural history and longitudinal cohorts between September 1, 2015 to November 1, 2023 were studied (Queen Square Research Ethics Committee 14/LO/1575) **(1)**. We obtained written informed consent from all participants who were also offered the opportunity to register for post-mortem brain donation at 1 of 4 UK brain banks (Queen Square [London], Cambridge, Oxford, and Manchester).

**CSF alpha-synuclein SAA analysis**

80/106 participants underwent baseline clinical assessment and lumbar puncture on the same day whilst the remaining participants had their lumbar puncture within three months of clinical assessment.

The alpha-synuclein seed amplification assay was performed in the Amprion Clinical Laboratory (CLIA ID No. 05D2209417; CAP No. 8168002) using a method validated for clinical use in accordance with Clinical Laboratory Improvement Amendment (CLIA) requirements. Each sample is analysed in triplicate in a 96-well plate using a reaction mixture comprised of 100mM PIPES pH 6.5, 0.5M NaCl, 0.1% sarkosyl, 10µM ThT, 0.3mg/mL recombinant alpha-synuclein, and 40µL CSF, in a final volume of 100µL. Two silicon nitride beads are included in each well, and positive and negative assay quality control samples are included on each plate. Plates are sealed with optical adhesive film, placed into the chamber of a BMG LABTECH FLUOstar Ω Microplate Reader, and incubated at 42°C with cycles of 1 min of shaking followed by 14 minutes of rest with fluorescence measured after every shaking cycle (excitation wavelength 440 nm, emission 490 nm). After a total incubation time of 20 hours, the maximum fluorescence for each well is determined and an algorithm applied to the triplicate determinations for each sample for result classification.
